# COVID-19 Vaccination among People Affected by Sexually Transmitted and Bloodborne Infections, Methamphetamine Use and their Intersection in Manitoba, Canada: A Retrospective Matched Cohort Analysis Using Population-Based Administrative Healthcare Data (2020-2022)

**DOI:** 10.64898/2026.05.18.26353507

**Authors:** Souradet Y. Shaw, Alyson Mahar, Kim Bailey, Mike Payne, Jason Kindrachuk, Christine Kelly, Kevin John Friesen, Charles N. Bernstein, Joss Reimer, Marissa L. Becker, Leigh M. McClarty, Derek Stein, Nathan C. Nickel

**Affiliations:** College of Community and Global Health, University of Manitoba; Department of Medical Microbiology and Infectious Diseases, University of Manitoba; School of Nursing, Queen’s University; Manitoba Centre for Health Policy, University of Manitoba; Nine Circles Community Health Centre; Department of Internal Medicine, University of Manitoba; Public Health Agency of Canada; Cadham Provincial Laboratory

## Abstract

**Objectives:** To examine COVID-19 vaccine uptake among people diagnosed with sexually transmitted and bloodborne infections (STBBI), people with healthcare-documented methamphetamine use, and those experiencing both exposures in Manitoba, Canada.

**Methods:** We conducted a retrospective matched-cohort study using linked population-based administrative healthcare, laboratory, and vaccination databases in Manitoba. Individuals aged ≥16 years with laboratory-confirmed chlamydia/gonorrhea (CT/NG), syphilis, HIV, and/or documented methamphetamine use during the four years prior to March 1, 2020 were included in eight exposed cohorts. Each cohort was matched to unexposed comparators on age, sex, geographic region, and income quintile. The primary outcome was receipt of ≥2 COVID-19 vaccine doses between December 1, 2020 and March 31, 2022. Poisson regression models estimated adjusted rate ratios (aRRs) and 95% confidence intervals (95% CIs) for vaccine uptake.

**Results:** Compared with matched comparators, most exposed cohorts were less likely to complete the COVID-19 primary vaccine series. Individuals in the Syphilis Only (aRR: 0.83, 95% CI: 0.80–0.86), Syphilis Plus (aRR: 0.77, 95% CI: 0.74–0.79), CT/NG Only (aRR: 0.91, 95% CI: 0.90–0.92), CT/NG Plus (aRR: 0.74, 95% CI: 0.72–0.77), Methamphetamine Only (aRR: 0.71, 95% CI: 0.68–0.73), and Methamphetamine + STBBI cohorts (aRR: 0.64, 95% CI: 0.62–0.67) had significantly lower vaccine uptake. The HIV Only cohort did not differ significantly from matched comparators (aRR: 0.97, 95% CI: 0.93–1.004). Lower uptake was concentrated among individuals living in lower-income areas.

**Conclusions:** Populations affected by STBBI, reported methamphetamine use, or both experienced significant inequities in COVID-19 vaccine uptake, particularly those with multiple STBBIs and concurrent substance use. Integrated vaccination approaches linked with HIV, harm reduction, and addiction services may improve vaccine equity during future public health emergencies.

**STRENGTHS AND LIMITATIONS OF THIS STUDY:**

- Population-based study using objective measures of COVID-19 vaccination.
- Regression models addressed confounding by age, geographic area, area-level income, healthcare utilization, and previous mental health and alcohol use disorder diagnoses.
- Does not capture individuals not interacting with healthcare system.
- Residual confounding can still be present, with no information on behavioural risk factors for STBBI, race/ethnicity, and individual-level socioeconomic measures.

## INTRODUCTION

The speed and scale at which SARS-CoV-2, the virus that causes coronavirus disease 2019 (COVID-19), exposed underlying inequities across global societies was unprecedented, with socio-economically and structurally disadvantaged populations experiencing disproportionately high rates of infection, severe disease, and mortality.^1^ The central Canadian province of Manitoba experienced the highest cumulative COVID-19 mortality rate nationally,^2^ highlighting systemic vulnerabilities, including overcrowded housing, precarious employment, and barriers to healthcare access. ^3^

The public health response to COVID-19 disrupted routine healthcare services, disproportionately impacting marginalized populations, including those at risk for sexually transmitted and bloodborne infections (STBBI).^4^ STBBI contribute substantially to health, social, and economic burdens globally,^5^ with chlamydia (CT) and gonorrhea (NG) among the most widely reported bacterial infections worldwide.^5^ Untreated STBBI are associated with a range of sequelae and complications, including infertility.^1^ ^5^ Like COVID-19, STBBI risk is shaped by structural factors including poverty, stigma, racial inequity, and social marginalization.^6^

In Manitoba, infectious syphilis, congenital syphilis, and HIV reached historically high levels before and during the pandemic.^7^ ^8^ Research from Manitoba demonstrated that among people newly diagnosed with HIV between 2018 and 2021, 62–76% reported methamphetamine use, more than half reported injection drug use, and houselessness was common.^7^ Methamphetamine use was strongly associated with a higher prevalence of multiple STBBI before and after HIV diagnosis, reflecting a syndemic of STBBI, substance use, and houselessness.^7^ Thus, STBBI diagnoses and methamphetamine use can be conceptualized as distinct but intersecting indicators of populations affected by a broader syndemic of infectious disease, substance use, socioeconomic disadvantage, and barriers to healthcare access.

Structural determinants driving STBBI and methamphetamine vulnerability also influence healthcare utilization, including vaccination. These inequities have also resulted in sub-optimal outcomes directly and indirectly related to the COVID-19 pandemic;^9^ for example, a US-based study showed those with a diagnosed substance use disorder were at 8 times the odds of having COVID-19.^10^ Studies have demonstrated sub-optimal COVID-19 vaccine uptake among those reporting methamphetamine use,^11^ while among people who inject drugs, uptake as low as 4% has been reported in some high-income countries.^12^ Although COVID-19 vaccine uptake has been found to be sub-optimal in people living with HIV (PLHIV),^13^ a scoping review found heterogeneity in vaccine hesitancy among PLHIV, with intersecting factors such as unemployment, trust in the medical system, sexual orientation, and relationship with healthcare providers contributing to vaccination decisions.^14^ A systematic review estimated uptake at 72% for North and South America, and factors such as younger age, unemployment, and lower education associated with lower uptake.^13^ One population-based study found comparatively high uptake of COVID-19 vaccines in a cohort of PLHIV;^15^ however, this study only examined those living with HIV, and not those with other STBBIs or substance users.

Thus, important gaps remain in understanding COVID-19 vaccine coverage among people affected by STBBI other than HIV, particularly where substance use and other forms of structural vulnerability intersect. Given the well-documented syndemic relationship between STBBI and methamphetamine use in Manitoba, we examined vaccine uptake across distinct populations affected by STBBI, healthcare-documented methamphetamine use, or both during the acute phase of the pandemic. Using linked population-based administrative healthcare data, we quantified differences in COVID-19 vaccine coverage across these populations to inform more equitable vaccination strategies during future public health emergencies.

## METHODS

### Population Setting, Data Sources, & Study Design

Manitoba is a centrally-located Canadian province with a population of 1.4 million people; almost 70% of the Manitoba population resides in the capital city of Winnipeg. Manitoba often has the highest rates of CT and NG nationally,^16^ and more recently reported the highest rates of infectious and congenital syphilis,^17^ and HIV.^18^ This study used a retrospective observational analysis, using a matched-cohort design, utilizing public health laboratory data from Cadham Provincial Laboratory (CPL) and the Public Health Information Management System (PHIMS), linked to routinely collected population-based administrative healthcare data housed at the Manitoba Centre for Health Policy (MCHP). Manitobans are provided universal healthcare through a single insurer (Manitoba Health), and all residents of Manitoba are assigned personal health identification numbers to track reimbursement to healthcare professionals. CPL provides the majority (>95%) of testing for chlamydia and gonorrhea, and all testing for syphilis and HIV in Manitoba. PHIMS tracks all vaccinations administered in Manitoba. Population-based physician claims, hospitalizations, pharmaceutical dispensations, and other routine social and healthcare data can be linked at MCHP, creating longitudinal healthcare records of individuals receiving care in Manitoba.^19^

### Cohort Creation, Outcomes, and Comparator Cohorts

STBBI cohorts were defined using CPL data, and included all individuals aged 16 years or older, and who had a positive laboratory test for CT/NG, syphilis, and/or HIV in the four-year period (as measured by specimen collection date) prior to the COVID-19 pandemic reaching Manitoba on March 1, 2020. Positive tests were determined as per CPL protocols;^20^ for CT/NG, this meant detection through nucleic acid tests in genitourinary specimens; for HIV, serological detection of HIV-1/HIV-2 antibodies or detection of nucleic acid through PCR; for syphilis, identification of *T. pallidum* by dark-field microscopy, fluorescent antibody, or detection of *T. pallidum* DNA by NAAT. For each STBBI, individuals were grouped into an “Only” cohort, meaning a positive test for only that pathogen in the four-year period, or a “Plus” (or co-infection) cohort, which included members testing positive for more than one of the pathogens. For example, the “HIV Only” cohort included only those with a positive HIV test recorded in the four years prior to March 1, 2020, while the “HIV Plus” cohort included those with a positive test for HIV, and a positive test for one or more of CT, NG, or syphilis in the same time period. In addition to creating cohorts of people with positive tests for STBBI, we also created a cohort of individuals who had contact with the healthcare system because of their use of methamphetamine. Based on the work of Nickel et al., these individuals were identified using a combination of International Classification of Diseases (ICD) codes in hospitalization and physician visits data, keyword searches in emergency department and fire and paramedic services data, and from diagnostic tests where methamphetamine was identified.^21^ Similar to the STBBI cohorts, the case definition was applied to the four years prior to March 1, 2020. In total, 8 cohorts were created: HIV Only, HIV Plus, CT/NG Only, CT/NG Plus, Syphilis Only, Syphilis Plus, Methamphetamine Only, and Methamphetamine + STBBI (which included those with STBBI diagnoses). For the purposes of this manuscript, we will refer to these 8 cohorts as the “exposed” cohorts.

Scrambled identification numbers were used to deterministically link members of each cohort to administrative healthcare data housed at MCHP, including information from PHIMS. The outcome for each cohort was the receipt of two or more doses (“primary series”) of a COVID-19 vaccine between December 1, 2020 to March 31, 2022. For the purposes of these analyses, all individuals had to have been continuously registered in Manitoba Health’s insurance registry from the start of the study (February 1, 2016), to the end of the study (March 31, 2022). Thus, anyone who left the province of Manitoba, or who died during the study period were excluded; less than 0.5% of our sample were excluded for these reasons. Individuals in each of the cohorts were matched (without replacement) to Manitobans with no history of a positive STBBI test, or evidence of methamphetamine use recorded in Manitoba during the study period at a 5:1 ratio based on sex, birth year, and forward sortation area (FSA, or the first 3 digits of the postal code), and income quintile as of February 1, 2020. Because of the large number of CT/NG cases, the matching ratio was 3:1. Thus, each cohort was matched to their own comparator cohort; we refer to the comparator cohorts as “unexposed”.

Area-level income quintiles were created using data from Statistics Canada and were defined using dissemination area and linked to individuals by their postal code; areas included in income quintile 1 (IQ1) were considered to have the lowest income levels, while income quintile 5 (IQ5) had the highest income levels. Rural and Urban income quintiles were combined for these analyses. Ambulatory physician visits in the year prior to February 1, 2020 were recorded and categorized as 0, 1-2, 3-4, and 5+ visits, and were treated as confounders in regression models. Other confounders included a binary indicator of any mental health diagnosis in the year prior to February 1, 2020, using established case-finding algorithms for psychotic disorders, mood/anxiety disorders, and attention deficit/hyperactivity disorder.^22^ Finally, and using the same one-year window, we created a binary indicator for alcohol use disorder.^23^ This project received ethics approval from the Human Research Ethics Board at the University of Manitoba (HS25742).

### Statistical Analyses

Descriptive statistics were used to describe cohorts by age group, sex, regional health authority, income quintile, physician visits in the year prior to the pandemic, and by COVID-19 vaccination status. Partially and fully (adjusted for physician visits, mental health diagnoses, and diagnosed alcohol use disorder, in addition to all matched variables) Poisson regression models examining the association between cohort membership (i.e., each “exposed” cohort and their “unexposed” comparator cohort) and outcomes were estimated for each of the 8 cohorts. Person-years (PY) was used as the offset variable, and partially and adjusted rate ratios (a/RRs) and 95% confidence intervals (95% CIs) were used to quantify the association, with a p-value of <.05 considered statistically significant. Person-years were used as an offset to account differences in follow-up time for individuals to receive their COVID-19 vaccinations; thus, a person receiving their COVID-19 dose(s) earlier on in the pandemic contributed less person-time, compared to a person receiving their vaccine later in the pandemic. As an additional analysis, equiplots (https://www.equidade.org/en/equiplot) were used to illustrate inequalities in vaccination in each exposed cohort, by income quintile.

### Patient and Public Involvement

No patient or public involvement in study design or conduct of the study. Study results have been disseminated to community organizations in Winnipeg.

## RESULTS

For the exposed cohorts, a total of 2,045 individuals were included in the Syphilis Only cohort, and 2,482 included in the Syphilis Plus cohort (Table 1). There were 995 and 377 individuals in the HIV Only and Plus cohorts, respectively; 21,984 and 65,034 in the CT/NG Only and Plus cohorts; and 2,832 and 2,232 in the Methamphetamine Only and Methamphetamine + STBBI cohorts. Table 1 shows the socio-demographic and COVID-19 related characteristics of each exposed cohort. Approximately 70% of the Syphilis Only cohort were less than 45 years of age, compared to 90% of the Syphilis Plus cohort. The difference in age distribution was mostly due to the Syphilis Plus cohort containing more individuals aged 16-24 years (28% vs. 11%). The difference in age between “Only” and “Plus” cohorts was true with each of the exposed cohorts, with the exception of the CT/NG cohort; 61% of the HIV Plus cohort were under the age of 44 years, compared to 42% of the HIV Only cohort, while 93% of the Methamphetamine + STBBI cohort were less than 45 year of age, compared to 81% of the Methamphetamine Only cohort. In contrast, the CT/NG Plus cohort contained more individuals aged between 25-44 years of age, compared to the CT/NG Only cohort (65% vs. 53%). The Syphilis Plus and Methamphetamine + STBBI cohorts were composed of more women, compared to the Syphilis and Methamphetamine Only cohorts (56% vs. 36% and 64% vs. 40%, respectively). With the exception of the HIV cohorts, each “Plus” (and for Methamphetamine, the + STBBI) cohort contained more individuals living in lowest income quintile areas, compared to their respective singly exposed cohort. For example, 44% of the CT/NG Plus cohort lived in the lowest income quintile areas, compared to 31% of the CT/NG Only cohort. Mental health diagnoses in the year prior to the pandemic ranged from 19.2% (HIV Only) to 50.4% (Meth Only); diagnosed alcohol use disorder was also highest in the Meth Only cohort (12.4%), while lowest in the CT/NG Only cohort (2.8%).

**Table 1.** Socio-Demographic and COVID-19-Related Characteristics, Exposed Cohorts, Sexually Transmitted and Bloodborne Infection and Methamphetamine Cohorts, Manitoba (2020-2022).

|  |  | Syphilis (n, %) |  | HIV (n, %) |  | Chlamydia /Gonorrhea (n, %) |  | Methamphetamine (n, %) |  |
| --- | --- | --- | --- | --- | --- | --- | --- | --- | --- |
|  |  | Syphilis Only (n=2,045) | Syphilis Plus (n=2,482) | HIV Only (n=995) | HIV Plus (n=377) | CT/NG Only (n=21,984) | CT/NG Plus (n=2,430) | Meth Only (n=2,832) | Meth + STBBI (n=2,232) |
| Age group (years) |  |  |  |  |  |  |  |  |  |
|  | 16-24 | 217 (10.6) | 692 (27.9) | 21 (2.1) | 20 (5.3) | 9,190 (41.8) | 693 (28.5) | 434 (15.3) | 552 (24.7) |
|  | 25-44 | 1,235 (60.4) | 1,548 (62.4) | 335 (39.6) | 208 (55.2) | 11,740 (53.4) | 1,573 (64.7) | 1,848 (65.3) | 1,528 (68.5) |
|  | 45-59 | 425 (20.8) | 196 (7.9) | 470 (43.1) | 122 (32.4) | 945 (4.3) | 136 (9.5) | 480 (16.9) | 141 (6.3) |
|  | 60+ | 168 (8.2) | 46 (1.9) | 169 (17.0) | 27 (7.2) | 109 (0.5) | 28 (2.0) | 70 (2.5) | 11 (0.05) |
| Sex |  |  |  |  |  |  |  |  |  |
|  | Male | 1,301 (63.6) | 1,109 (44.4) | 627 (63.0) | 253 (57.1) | 8,789 (40.0) | 1,028 (42.3) | 1,691 (59.7) | 798 (35.8) |
|  | Female | 744 (36.4) | 1,379 (55.6) | 368 (37.0) | 124 (32.9) | 13,195 (60.0) | 1,402 (57.7) | 1,141 (40.3) | 1,434 (64.2) |
| Regional Health Authority |  |  |  |  |  |  |  |  |  |
|  | Interlake-Eastern | 146 (7.1) | 157 (6.3) | 57 (5.7) | 21 (5.6) | 2,132 (9.7) | 157 (6.5) | 181 (6.4) | 162 (7.3) |
|  | Northern | 489 (23.9) | 647 (26.1) | 44 (4.4) | 22 (5.8) | 4,727 (21.5) | 648 (26.7) | 140 (4.9) | 169 (7.6) |
|  | Southern | 111 (5.4) | 78 (3.1) | 45 (4.5) | s | 1,614 (7.3) | 75 (3.1) | 140 (4.9) | 78 (3.5) |
|  | Prairie Mountain | 105 (5.1) | 93 (3.7) | 54 (5.4) | 15 (4.0) | 2,172 (9.9) | 91 (3.7) | 273 (9.6) | 134 (6.0) |
|  | Winnipeg | 1,194 (58.4) | 1,507 (60.7) | 795 (79.9) | 319 (84.6) | 11,339 (51.6) | 1,459 (60.0) | 2,098 (74.1) | 1,689 (75.7) |
| Income Quintile |  |  |  |  |  |  |  |  |  |
|  | Q1 (Lowest) | 787 (38.5) | 1,100 (44.3) | 534 (53.7) | 203 (53.8) | 6,839 (31.0) | 1,073 (44.2) | 1,399 (49.4) | 1,245 (55.8) |
|  | Q2 | 581 (28.4) | 776 (31.3) | 179 (18.0) | 70 (18.6) | 6,315 (28.7) | 765 (31.5) | 611 (21.6) | 503 (22.5) |
|  | Q3 | 265 (13.0) | 218 (8.8) | 127 (12.8) | 43 (11.4) | 3,527 (16.0) | 208 (8.6) | 369 (13.0) | 190 (8.5) |
|  | Q4 | 270 (13.2) | 273 (11.0) | 98 (9.8) | 40 (11.9) | 2,980 (13.6) | 273 (11.2) | 290 (10.2) | 206 (9.2) |
|  | Q5 (Highest) | 142 (6.9) | 115 (4.6) | 57 (5.7) | 21 (5.6) | 2,323 (10.6) | 111 (5.6) | 163 (5.8) | 88 (3.9) |
| Physician Visits, 1 |  |  |  |  |  |  |  |  |  |
| Year Prior to COVID- |  |  |  |  |  |  |  |  |  |
| 19 Pandemic |  |  |  |  |  |  |  |  |  |
|  | 0 | 270 (13.2) | 384 (15.5) | 40 (4.0) | 16 (4.2) | 3,505 (15.9) | 383 (15.8) | 244 (8.6) | 177 (7.9) |
|  | 1-2 | 205 (10.0) | 286 (11.5) | 41 (4.1) | 16 (4.2) | 2,326 (10.6) | 287 (11.8) | 182 (6.4) | 161 (7.2) |
|  | 2-4 | 435 (21.3) | 552 (22.2) | 187 (18.8) | 84 (22.3) | 5,438 (24.7) | 540 (22.2) | 387 (13.7) | 351 (15.7) |
|  | 5+ | 1,135 (55.5) | 1,260 (50.8) | 727 (73.1) | 261 (69.2) | 10,715 (48.7) | 1,220 (50.2) | 2,019 (71.3) | 1,543 (69.1) |
| Mental Health |  |  |  |  |  |  |  |  |  |
| Diagnosis (1 year prior |  |  |  |  |  |  |  |  |  |
| to pandemic) |  |  |  |  |  |  |  |  |  |
|  | No | 1,622 (79.3) | 1,932 (77.8) | 804 (80.8) | 286 (75.9) | 17,693 (80.5) | 1,888 (77.7) | 1,404 (49.6) | 1,279 (57.3) |
|  | Yes | 423 (20.7) | 550 (22.2) | 191 (19.2) | 91 (24.1) | 4,291 (19.5) | 542 (22.3) | 1,428 (50.4) | 953 (42.7) |
| Alcohol Use Disorder |  |  |  |  |  |  |  |  |  |
| Diagnosis (1 year prior |  |  |  |  |  |  |  |  |  |
| to pandemic) |  |  |  |  |  |  |  |  |  |
|  | No | 1,935 (94.6) | 2,337 (94.2) | 957 (96.2) | 357 (94.7) | 21,366 (97.2) | 2,283 (94.0) | 2,480 (87.6) | 1,962 (87.9) |
|  | Yes | 110 (5.4) | 145 (5.8) | 38 (3.8) | 20 (5.3) | 618 (2.8) | 147 (6.1) | 352 (12.4) | 270 (12.1) |

Apart from the HIV Only cohort, those exposed were less likely to have received the primary series of vaccinations than their matched comparator cohort (Table 2 & Figure 1). For example, 72.3% [vaccination rate: 480.7 (95% CI: 456.8-505.9) per 1,000 PY] of individuals in the Syphilis Only cohort had received their primary series, compared to 80.6% of their matched cohort [557.0 (95% CI: 545.1-569.2) per 1,000 PY]. Similarly, 68.1% [vaccination rate: 428.1 (95% CI: 408.2-449.0) per 1,000 PY] of the Syphilis Plus cohort received their complete primary series, compared to 79.7% [544.1 (95% CI: 533.5-554.9) per 1,000 PY] of their matched cohort. In comparison, 84.8% [597.0 (95%CI: 558.0-638.6) per 1,000 PY] of the HIV Only cohort received their primary series, compared to 82.5% [577.3 (95% CI: 559.9-595.2) per 1,000 PY] of the matched cohort. Of note, the proportion vaccinated in the HIV Plus cohort was 75.6% [511.2 (95% CI: 455.2-574.1 per 1,000 PY), lower than their matched cohort [79.6%, or 547.6 (95% CI: 520.1-576.0) per 1,000 PY].

**Figure 1.**
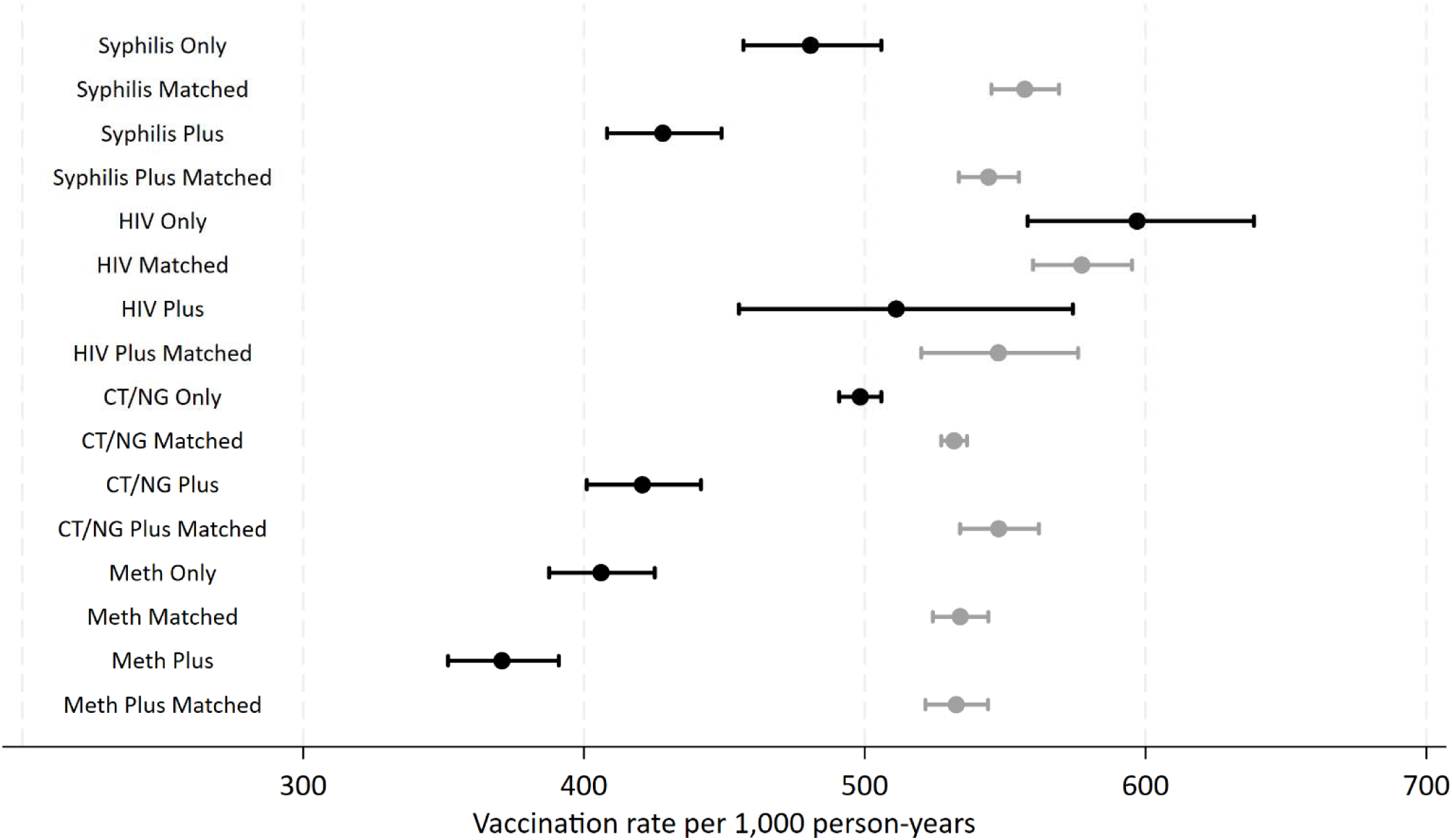
Vaccination Rate and 95% Confidence Intervals (per 1,000 person-years) for COVID-19 Vaccine Uptake (2+ Doses), by Sexually Transmitted and Bloodborne Infection (STBBI) and Methamphetamine Cohorts, Manitoba (2020-2022)^a^. ^a^‘Only’: Only that infection/condition detected in administrative healthcare and laboratory data; ‘Plus’: co-infection/co-morbidity with at least one other STBBI [incl. syphilis, chlamydia/gonorrhea (CT/NG), and/or HIV] in the four years prior to March, 2020

**Table 2.** Total Number, Vaccination Rate and 95% Confidence Intervals (per 1,000 person-years) of Exposed Cohorts and their Matched Comparators^a^, Sexually Transmitted and Bloodborne Infection (STBBI) and Methamphetamine Cohorts, Manitoba (2020-2022)

| **Cohort** | **N** | **2+ COVID-19**<br>**Vaccinations (n, %)** | **Vaccination Rate per 1,000**<br>**person-years (95% CI)** |
| --- | --- | --- | --- |
| Syphilis Only | 2,045 | 1,479 (72.3) | 480.7 (456.8-505.9) |
| *Matched cohort* | 10,225 | 8,237 (80.6) | 557.0 (545.1-569.2) |
| Syphilis Plus | 2,482 | 1,691 (68.1) | 428.1 (408.2-449.0) |
| *Matched cohort* | 12,410 | 9,888 (79.7) | 544.1 (533.5-554.9) |
| HIV Only | 995 | 844 (84.8) | 597.0 (558.0-638.6) |
| *Matched cohort* | 4,975 | 4,104 (82.5) | 577.3 (559.9-595.2) |
| HIV Plus | 377 | 285 (75.6) | 511.2 (455.2-574.1) |
| *Matched cohort* | 1,885 | 1,501 (79.6) | 547.6 (520.1-576.0) |
| Chlamydia/Gonorrhea Only | 21,984 | 16,817 (76.5) | 498.4 (490.9-505.9) |
| *Matched cohort* | 65,034 | 51,056 (78.5) | 531.8 (527.2-536.4) |
| Chlamydia/Gonorrhea Plus | 2,430 | 1,640 (67.5) | 420.8 (401.0-441.7) |
| *Matched cohort* | 7,290 | 5,828 (80.0) | 547.7 (533.9-562.0) |
| Methamphetamine Only | 2,832 | 1,825 (64.4) | 406.1 (387.9-425.2) |
| *Matched cohort* | 14,160 | 11,128 (78.6) | 534.0 (524.2-544.0) |
| Methamphetamine + STBBI | 2,232 | 1,366 (61.2) | 370.8 (351.6-391.0) |
| *Matched cohort* | 11,160 | 8,747 (78.4) | 532.6 (521.6-543.9) |

<sup>a</sup>Matched by age, sex, income quintile and regional health authority at a 5:1 ratio, with the exception of the chlamydia/gonorrhea cohort, which was matched at a 3:1 ratio

Table 3 contains the results from Poisson regression models examining the association between membership in exposed cohorts and the likelihood of receiving the primary series of COVID-19 vaccinations. In fully adjusted models, and compared to their matched unexposed cohort, those belonging to the Syphilis Only cohort were 17% less likely (aRR: 0.83, 95% CI: 0.80-0.86) to receive their primary series; this difference increased to 23% (aRR: 0.77, 95% CI: 0.74-0.79) for the Syphilis Plus cohort. Similarly, those in the CT/NG Only cohort were 9% less likely (aRR: 0.91, 95% CI: 0.90-0.92) to receive their primary series, while those in the CT/NG Plus cohort were 26% less likely (aRR: 0.74, 95% CI: 0.72-0.77), compared to their respective matched cohort. The aRRs were 0.71 (95% CI: 0.68-0.73) and 0.64 (95% CI: 0.62-0.67) for the Methamphetamine Only and Methamphetamine + STBBI cohorts for primary series vaccinations, respectively, compared to their matched cohort. For HIV, while those in the HIV Plus cohort were significantly less likely to have received their primary series (aRR: 0.87, 95% CI: 0.81-0.94), this association was not statistically significant for the HIV Only cohort (aRR: 0.97, 95% CI: 0.93-1.004).

**Table 3.** Rate Ratios (RRs) and 95% Confidence Intervals (95% CI) from Partial and Full^a^ Poisson Regression Models Examining the Association between 2+ COVID-19 Vaccinations of Exposed Cohorts and their Matched Comparators^b^, Sexually Transmitted and Bloodborne Infection (STBBI) and Methamphetamine Cohorts, Manitoba (2020-2022).

|  |  | 2+ COVID-19 Vaccinations<br>[RR (95%CI)] |  |
| --- | --- | --- | --- |
|  |  | Partial | Full<sup>a</sup> |
| *Syphilis* |  |  |  |
|  | Syphilis Only<sup>c</sup> | 0.86 (0.83-0.90) | 0.83 (0.80-0.86) |
|  | Syphilis Plus<sup>d</sup> | 0.79 (0.76-0.82) | 0.77 (0.74-0.79) |
| *HIV* |  |  |  |
|  | HIV Only<sup>c</sup> | 1.03 (0.996-1.07) | 0.97 (0.93-1.004) |
|  | HIV Plus<sup>d</sup> | 0.94 (0.86-1.01) | 0.87 (0.81-0.94) |
| *Chlamydia(CT)/Gonorrhea (NG)* |  |  |  |
|  | CT/NG Only<sup>c</sup> | 0.94 (0.93-0.95) | 0.91 (0.90-0.92) |
|  | CT/NG Plus<sup>d</sup> | 0.77 (0.74-0.80) | 0.74 (0.72-0.77) |
| *Methamphetamine (Meth)* |  |  |  |
|  | Meth Only<sup>c</sup> | 0.76 (0.74-0.79) | 0.71 (0.68-0.73) |
|  | Meth + STBBI<sup>d</sup> | 0.70 (0.67-0.73) | 0.64 (0.62-0.67) |

<sup>a</sup>Full models include number of physician visits in the year prior to the COVID-19 pandemic, categorized as 0, 1-2, 3-4, 5+ visits; and binary indicators for mental health diagnoses and diagnosed alcohol use disorder.
<sup>b</sup>Matched by age, sex, income quintile and regional health authority at a 5:1 ratio, with the exception of the chlamydia/gonorrhea cohort, which was matched at a 3:1 ratio
<sup>c</sup>No other STBBI co-infection; <sup>d</sup>Multiple infections with at least one other STBBI (incl. syphilis, CT/NG, and/or HIV) in the four years prior to March, 2020

Figure 2 contains a visualization of inequalities in the distribution of COVID-19 vaccinations, by income quintile for all 8 exposed cohorts, and for Manitoba overall. In almost all cohorts, those living in lowest income quintile areas (IQ1) substantially trailed those living in areas with higher income quintiles for primary series vaccinations. For example, in the Syphilis Only cohort, approximately 67% of those living in the lowest income communities received their primary series, compared to 74% in the next income quintile, for a risk difference of 7%. The proportion living in the lowest income communities receiving their primary series was even lower in the Syphilis Plus cohort, at 61%.

**Figure 2.**
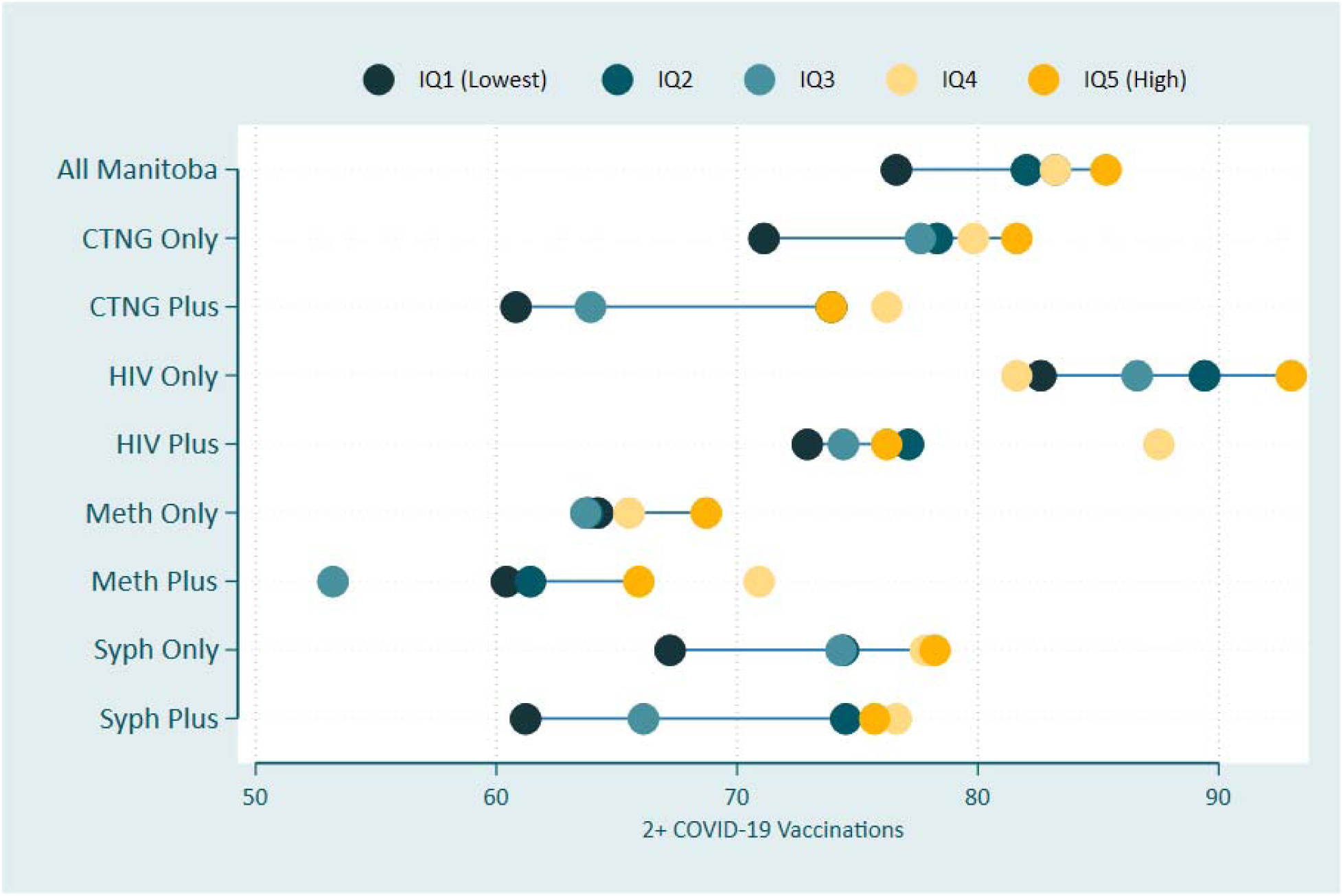
Equiplots Illustrating COVID-19 Vaccine Uptake (2+ Doses), by Income Quintile, Sexually Transmitted and Bloodborne Infections (STBBI) and Methamphetamine Cohorts, Manitoba (2020-2022)^a^. ^a^‘Only’: Only that infection/condition detected in administrative healthcare and laboratory data; ‘Plus’: co-infection/co-morbidity with at least one other STBBI (incl. syphilis, CT/NG, and/or HIV) in the four years prior to March, 2020

## DISCUSSION

Our findings demonstrate that COVID-19 vaccine uptake was lower across several distinct populations affected by STBBI, healthcare-documented methamphetamine use, or both, compared with matched comparator cohorts. However, the magnitude of this disparity varied. Uptake in the HIV Only cohort was comparable to that of matched comparators, whereas it was modestly lower among people with CT/NG alone and substantially lower among those with multiple STBBI diagnoses or healthcare-documented methamphetamine use. The greatest disparity occurred among individuals with both methamphetamine use and STBBI, who were approximately 36% less likely to complete the primary vaccine series.

Although Canada achieved high overall COVID-19 vaccine coverage,^24^ substantial inequities persisted among marginalized populations.^14^ ^25^ Our findings are consistent with studies documenting suboptimal vaccination among people who use drugs and those experiencing housing instability.^14^ In Vancouver’s inner city, fewer than half of participants reporting unstable housing and injection drug use had completed the primary vaccine series.^26^ Similarly, a study among people who inject drugs in San Diego found that only 39% had received at least one vaccine dose by the end of March 2022.^27^ In New York City, vaccine hesitancy among people who inject drugs was associated with limited prior vaccination and negative attitudes toward vaccines.^28^ Together, these findings suggest that barriers extend beyond vaccine availability and may include unstable living conditions, limited healthcare engagement, mistrust, and competing social and health priorities.

The HIV Only cohort represented an important exception; vaccine uptake in this group was comparable to matched controls, consistent with findings from Ontario, Canada.^15^ In their retrospective matched-cohort study of people living with HIV in Ontario, Freitas et al. found uptake of 2+ doses of the COVID-19 vaccine was at 85% for those with HIV, and 83% for those not living with HIV;^15^ exactly the proportions we report for the HIV Only and its matched cohort. In their scoping review, Newman et al. found that vaccine hesitancy among people living with HIV was dependent on a number of factors, some of which were protective against hesitancy.^14^ These included individual-level factors such as the perceived threat of COVID-19 on their health, and level of engagement with healthcare practitioners.^14^ Manitoba has a centralized program for people living with HIV – the Manitoba HIV Program; this program has prioritized engagement and sustaining care for people living with HIV. During the acute phase of the pandemic, the Manitoba HIV Program pivoted to virtual care models, while also prioritizing COVID-19 vaccines when they became available for people living with HIV. Co-location of COVID-19 vaccination service with other services related to injection drug use has been found to be successful from the vaccination uptake perspective, with one study showing PWID using co-located services having higher-than-average (94%) uptake of 2+ doses of the vaccine.^29^

Our observation that STBBI co-infected cohorts experienced the lowest vaccine uptake is novel. Previous Manitoba research has shown that STBBI co-infections are associated with younger age, inner-city residence, ^30^ repeat infections, ^31^ and higher social deprivation.^32^ These same structural determinants likely contribute to inequities in vaccination. Understanding barriers and facilitators to vaccine uptake in these populations may help inform future vaccination strategies, including for emerging STBBI vaccines.

Strengths of this study include the use of linked population-based datasets, objective vaccination measures, and the ability to define co-infected STBBI cohorts. We also adjusted for healthcare utilization, mental health diagnoses, and diagnosed alcohol use disorders. Limitations include the inability to capture individuals not interacting with the healthcare system; however, we would expect that this would bias our results even more positively; that is, a greater discrepancy in vaccine uptake would be shown between people with STBBI who do not regularly access healthcare and a comparable cohort. We also lacked detailed information regarding behavioral risk factors for STBBI, individual-level socioeconomic measures, and race/ethnicity data, thus residual confounding may still be present.

COVID-19 vaccine uptake among people living with HIV without other STBBI diagnoses did not differ significantly from matched comparators, suggesting that sustained engagement in specialized HIV care may support equitable vaccine delivery. In contrast, individuals with multiple STBBI diagnoses and methamphetamine use were substantially less likely to complete the primary vaccine series. Understanding how infection status, substance use, and socioeconomic inequities intersect is essential for designing equitable vaccination strategies during future public health emergencies.

## Data Availability

Data are protected by provincial legislation and are not freely available.

## Acknowledgements

*“The authors acknowledge the Manitoba Centre for Health Policy for use of data contained in the Manitoba Population Research Data Repository under project #55932 (PHRPC#P2022-117). The results and conclusions are those of the authors and no official endorsement by the Manitoba Centre for Health Policy, Manitoba Health, or other data providers is intended or should be inferred. Data used in this study are from the Manitoba Population Research Data Repository housed at the Manitoba Centre for Health Policy, University of Manitoba and were derived from data provided by Manitoba Health.”*

